# Socioeconomic inequalities in sport participation: pattern per sport and time trends

**DOI:** 10.1101/2022.11.18.22282493

**Authors:** Viviane Richard, Giovanni Piumatti, Nick Pullen, Elsa Lorthe, Idris Guessous, Nicola Cantoreggi, Silvia Stringhini

## Abstract

**Background:** Sport participation is an important component of a healthy lifestyle and is known to be more common among privileged individuals. However, few studies have examined socio-demographic patterns of participation by type of activity. This study aims at quantifying socio-economic inequalities in sport participation by sport type, and to analyse their trend over 15 years.

**Methods:** We used 2005-2019 data from the Bus Santé study, a yearly population-based cross-sectional survey of Geneva adults. Sport participation was defined as reporting at least one sporting activity over the previous week; educational level, household income and occupational position were used as indicators of socio-economic position. Socio-economic inequalities in sport participation, and their trend over time, were examined using the relative and slope indexes of inequality (RII/SII).

**Results:** Out of 7769 participants (50.1% women, mean age 46 years old), 60% participated in a sporting activity. Results showed that the higher the socioeconomic circumstances, the higher the sport participation (RII=1.78; 95% Confidence Interval (CI): 1.64-1.92; SII=0.33; 95%CI: 0.29-0.37 for education). Relative inequalities varied per sport e.g., 0.68 (95%CI: 0.44-1.07) in football participation and 4.25 (95%CI: 2.68-6.75) in tennis/badminton participation for education. Yearly absolute inequalities in sport participation tended to increase between 2005 and 2019 for household income (β=0.01; P-value*=*0.024).

**Conclusion:** We observed strong socio-economic inequalities in sport participation in Geneva, with different magnitude depending on the type of sport. These inequalities seemed to increase over the 2005-2019 period. Our results call for tailored measures to promote the participation of socially disadvantaged populations in sporting activities.

**KEY MESSAGES:**

- Sport participation is associated with higher socioeconomic conditions. Research on the patterning of inequalities per sport type and on their trend over time remains scarce.
- There are high growing socioeconomic inequalities in sport participation in Geneva. Inequalities in sport participation are heterogeneous and lower in sports practised in group.
- Tailored sport promotion measures are needed for disadvantaged populations.

## 1. INTRODUCTION

Physical activity has important health benefits[1–4] and the World Health Organization recommends that adults engage in at least 150 minutes of moderate-intensity physical activity per week, 75 minutes of vigorous physical activity, or a combination of both[5]. However, in high-income countries, a meaningful share of the population does not participate in sufficient physical activity to meet these guidelines[6]. This is thought to contribute to the burden of non-communicable diseases[7].

As a specific type of physical activity, sport can represent an interesting way of balancing the lack of movement of many daily occupations, with additional benefits such as a positive impact on mental health and social cohesion[3,8]. The determinants of sport participation have been widely studied and favourable socio-economic conditions are consistently associated with higher sport participation[9–11]. Indeed, privileged individuals may have more psychosocial, financial and neighbourhood resources, as well as a higher health literacy to establish healthy behaviours[9–11].

In most studies examining the social patterning of physical activity, sport participation is analysed globally, whereas different patterns could exist as per sport type[12]. For example, United-States and Australian studies found that privileged socio-economic conditions are associated with higher general sport participation, but that this relationship is reversed for team sports participation[12,13]. A French study obtained similar results, with higher education and income being associated with individual sport participation, but not with team sport participation[14]. These differences are usually explained by socio-cultural factors or by the fact that group activities are more affordable compared to other sports. Very few epidemiological studies stratified the association between socio-economic conditions and sport participation by type of sport, while a more detailed understanding of this relationship would help in designing tailored and effective promotion strategies, especially for disadvantaged populations.

The evolution of the association between socio-economic conditions and sport participation also remains unclear, as some studies report increasing inequalities over time[15,16], while others do not find significant trends[17–20]. In sum, the evolution of social inequalities in sport participation seems context- and time-dependent. Furthermore, gender-specific analyses are rare and inconsistent[15,17,19], and recent data is lacking, with very few estimates after 2012[16,20].

The goal of the present study is to evaluate the association between socio-economic conditions and participation in different sports; and to analyse its evolution over a 15 years study period in the canton of Geneva, Switzerland.

## 2. METHODS

### Study population

Data came from the Bus Santé study, an ongoing yearly population-based cross-sectional survey conducted since 1993 in Geneva, an urban Swiss canton[21]. Every year, an age- and sex-stratified random sample of about 1000 non-institutionalized residents aged 20-75 years (35-75 years before 2012) provided by the local authorities was recruited through an initial invitation letter. Non-respondents were contacted with up to seven phone calls and two additional letters. Participants completed socio-demographic, lifestyle and health questionnaires, and attended a medical check-up, during which questionnaires were verified by a trained research nurse[21]. The Bus Santé study was approved by the Institute of Ethics Committee of the University of Geneva. All participants signed a written informed consent.

In the present study, we included participants of the Bus Santé surveys between 2005 and 2019. Average annual participation rate (number of participants/number of eligible invited persons) was 36.8% [range: 29.0-47.0%]. These estimates are conservative since it was not possible to identify if participants unreachable by phone [annual range: 18.1-54.9%] did not want to participate or did not receive the invitation letters.

We further selected participants who were physically capable of engaging in a sporting activity; physical incapacity being defined as an affirmative answer to either of the questions: “*Over the 4 last weeks, did you have difficulties showering or bathing, getting dressed, getting in/up from your bed or a chair, using the toilets or eating?*” and “*Over the 4 last weeks, did you have difficulties shopping or doing routine household chores?*”. Since individuals aged 20-34 years old were only recruited from 2012 onwards in the Bus Santé study, participants were divided into two subgroups for the following analyses: 1) participants aged 20-75 years old recruited between 2012 and 2019 for the analyses by sport category, and 2) participants aged 35-75 years old recruited between 2005 and 2019 for the time trend analyses (Supplementary 1).

## Measures

### Outcome

The main outcome was sport participation, overall, by sport category and by sport type, reported in a dedicated subsection of a validated physical activity frequency questionnaire (PAFQ)[22], and defined as the participation in at least one sporting activity in the week preceding the Bus Santé appointment. The 17 different sport types proposed in the PAFQ were grouped into five categories based on the need for facilities, equipment and/or partners: 1) Outdoor individual sports, requiring no specific facilities and minimal equipment (running, brisk walking, racing bicycle); 2) Individual sports requiring access to facilities but minimal equipment (strength training/weight lifting, swimming, ice-/roller-skating); 3) Racket sports and martial arts, requiring access to facilities, some equipment and partners (judo/karate, tennis/badminton, squash); 4) Group sports, requiring access to facilities and minimal equipment, usually practiced in group or team sessions (dance, European football, handball, gymnastics); 5) Special sports requiring longer trips to reach specific places or facilities and significant equipment (golf, downhill/water skiing, cross-country skiing, diving). Participants could report participation in more than one sport type or category.

### Explanatory variables

Socio-economic conditions were separately measured with education, household income and occupational position. Based on the main milestones of the Swiss education system, the educational level was divided into lower secondary (ISCED 2011 levels 0-2), upper secondary (ISCED 2011 levels 3-4) and tertiary (ISCED 2011 levels 5-8)[23]. Categories of gross monthly household income were adjusted by the number of people living in the household using the OECD-modified scale[24]. The result was split into four categories: <3000 CHF, 3000-4999 CHF, 5000-6999 CHF and 7000 CHF. Finally, participants reported their current occupational position in one of the following four categories: manual worker, manual self-employed worker, non-manual worker, and non-manual manager.

### Covariates

The following demographic and health characteristics were considered as covariates: age at the moment of the interview, sex, and country of birth (Switzerland, Southern Europe, Western Europe, Eastern and South-Eastern Europe, Other), as well as health indicators such as self-reported health (very good, good, medium, poor, very poor), body mass index (BMI), active smoking and presence of a chronic disease (diabetes, hypertension, or cardiovascular disease).

## Statistical analyses

Socio-economic inequalities in sport participation were estimated using the relative and slope indexes of inequality (RII and SII). These regression-based measures summarise the outcome difference between the socio-economic extremes, while taking intermediate categories into account[25]. The RII evaluates the relative difference: a RII of 1.1 is interpreted as a 10% higher outcome prevalence in the most privileged socio-economic group compared with the least privileged one. The SII measures the absolute difference: a SII of 0.1 means that the prevalence of the outcome is 10 percentage points higher in the most privileged socio-economic group than in the least privileged one.

To compute these indexes, each category of the ordinal socio-economic variables was translated into a numerical rank equal to the proportion of participants with lower socio-economic conditions, and added as the independent variable in the models. The SII was based on linear regressions, while the RII was computed with generalized linear models following a quasi-Poisson distribution; robust standard errors were calculated. A first minimal model was adjusted for potential demographic confounders such as age, sex, an interaction between age and sex, and country of birth; a second full model further included the above-defined health factors to disentangle the effect of the health status in the association between socio-economic conditions and sport participation.

To evaluate the evolution of inequalities over time between 2005 and 2019, the participants’ visit year was added in the above-described models, both as a main effect and as an interaction term with all other covariates. The coefficient of the interaction between the socio-economic rank and the year (β) was used to give an estimation of the time trend in inequalities. A sensitivity analysis was performed by removing years that may overly influence the regression.

Complete case analyses were performed by excluding observations with missing data on the examined variables. Analyses per sport category and of trends over time were performed on the entire resulting sample, as well as with a sex stratification. All analyses were performed with R-4.0.3 and significance level was set to 5%; p-values of descriptive analyses were adjusted for multiple comparisons with the Bonferroni method.

## 3. RESULTS

Of the 8425 individuals taking part in the survey between 2012 and 2019, we excluded 656 who were physically unable to engage in sporting activities (Supplementary 1). Therefore, the study population for the main analyses consisted of 7769 participants, with a mean age of 46 years old (SD: 14.2 years), 50.1% being women (Table 1). A total of 4660 (60.0%) participants reported engaging in at least one sporting activity over the last week. Individual outdoor sports were the most practised, reported by 2368 (30.5%) participants, while racket sports and martial arts were only practised by 490 (6.3%) participants. Sport participation was more common among younger participants, born in Switzerland or in Western Europe, with a good self-reported health, a normal BMI, no chronic disease, no smoking, and higher socio-economic conditions (*P*<0.001; Table 1).

**Table 1.**
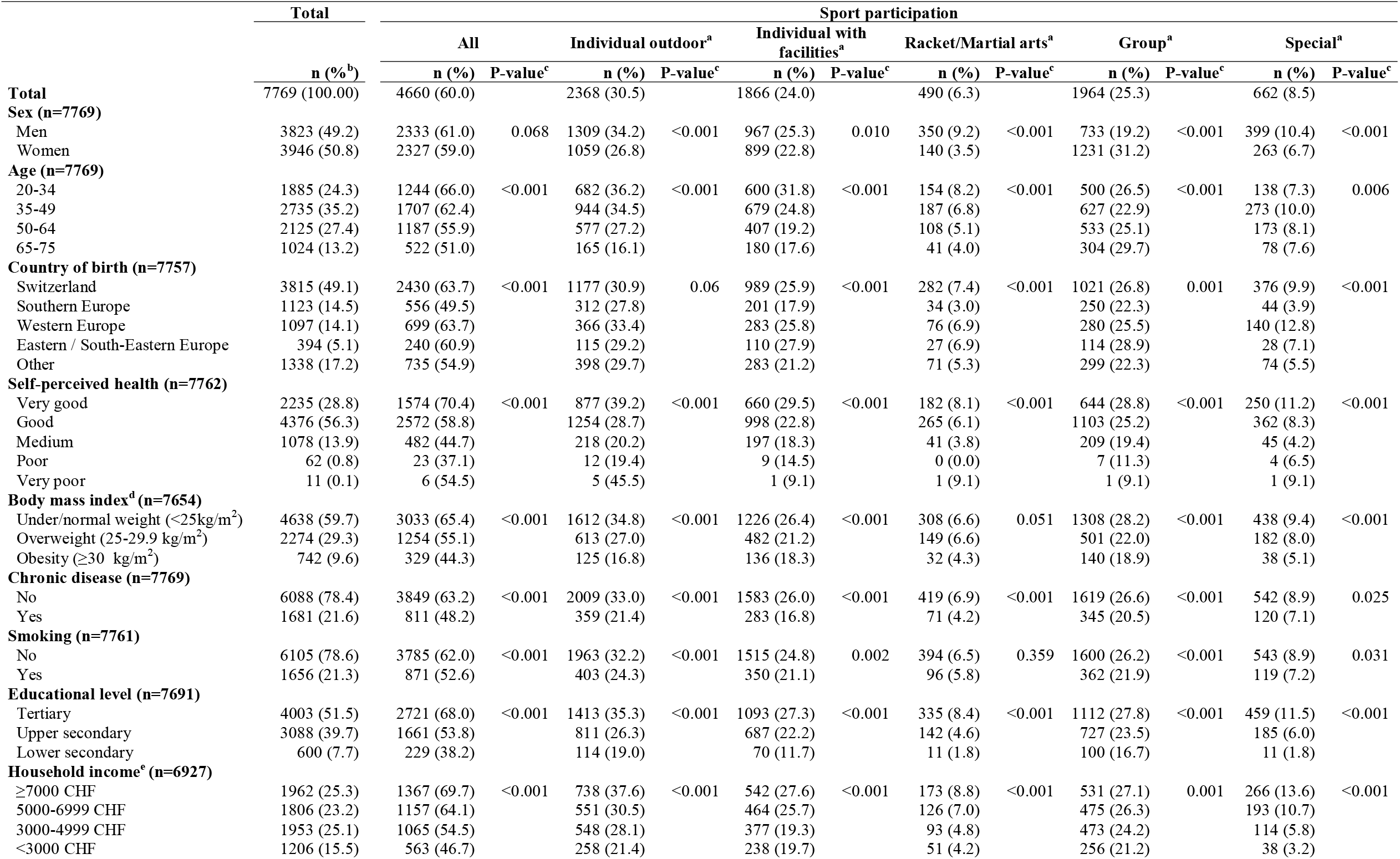
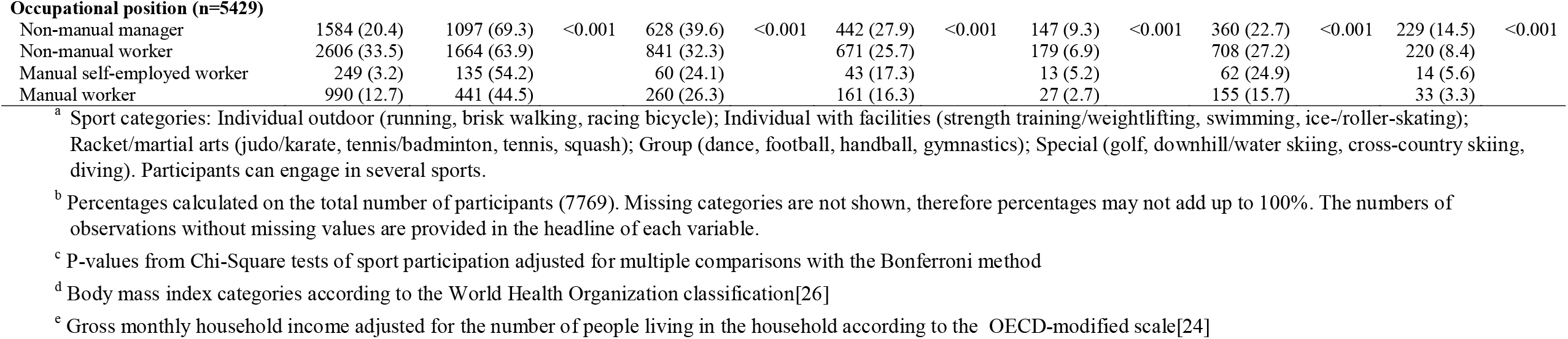
Demographic and health characteristics according to participation in sport categories, among adults aged 20 to 75 years, 2012-2019, Geneva

|  | Total | Sport participation |  |  |  |  |  |  |  |  |  |  |  |
| --- | --- | --- | --- | --- | --- | --- | --- | --- | --- | --- | --- | --- | --- |
|  | n (%)<sup>b</sup> | All |  | Individual outdoor<sup>a</sup> |  | Individual with facilities<sup>a</sup> |  | Racket/Martial arts<sup>a</sup> |  | Group<sup>a</sup> |  | Special<sup>a</sup> |  |
|  |  | n (%) | P-value<sup>c</sup> | n (%) | P-value<sup>c</sup> | n (%) | P-value<sup>c</sup> | n (%) | P-value<sup>c</sup> | n (%) | P-value<sup>c</sup> | n (%) | P-value<sup>c</sup> |
| **Total** | 7769 (100.00) | 4660 (60.0) |  | 2368 (30.5) |  | 1866 (24.0) |  | 490 (6.3) |  | 1964 (25.3) |  | 662 (8.5) |  |
| **Sex (n=7769)** |  |  |  |  |  |  |  |  |  |  |  |  |  |
| Men | 3823 (49.2) | 2333 (61.0) | 0.068 | 1309 (34.2) | <0.001 | 967 (25.3) | 0.010 | 350 (9.2) | <0.001 | 733 (19.2) | <0.001 | 399 (10.4) | <0.001 |
| Women | 3946 (50.8) | 2327 (59.0) |  | 1059 (26.8) |  | 899 (22.8) |  | 140 (3.5) |  | 1231 (31.2) |  | 263 (6.7) |  |
| **Age (n=7769)** |  |  |  |  |  |  |  |  |  |  |  |  |  |
| 20-34 | 1885 (24.3) | 1244 (66.0) | <0.001 | 682 (36.2) | <0.001 | 600 (31.8) | <0.001 | 154 (8.2) | <0.001 | 500 (26.5) | <0.001 | 138 (7.3) | 0.006 |
| 35-49 | 2735 (35.2) | 1707 (62.4) |  | 944 (34.5) |  | 679 (24.8) |  | 187 (6.8) |  | 627 (22.9) |  | 273 (10.0) |  |
| 50-64 | 2125 (27.4) | 1187 (55.9) |  | 577 (27.2) |  | 407 (19.2) |  | 108 (5.1) |  | 533 (25.1) |  | 173 (8.1) |  |
| 65-75 | 1024 (13.2) | 522 (51.0) |  | 165 (16.1) |  | 180 (17.6) |  | 41 (4.0) |  | 304 (29.7) |  | 78 (7.6) |  |
| **Country of birth (n=7757)** |  |  |  |  |  |  |  |  |  |  |  |  |  |
| Switzerland | 3815 (49.1) | 2430 (63.7) | <0.001 | 1177 (30.9) | 0.06 | 989 (25.9) | <0.001 | 282 (7.4) | <0.001 | 1021 (26.8) | 0.001 | 376 (9.9) | <0.001 |
| Southern Europe | 1123 (14.5) | 556 (49.5) |  | 312 (27.8) |  | 201 (17.9) |  | 34 (3.0) |  | 250 (22.3) |  | 44 (3.9) |  |
| Western Europe | 1097 (14.1) | 699 (63.7) |  | 366 (33.4) |  | 283 (25.8) |  | 76 (6.9) |  | 280 (25.5) |  | 140 (12.8) |  |
| Eastern / South-Eastern Europe | 394 (5.1) | 240 (60.9) |  | 115 (29.2) |  | 110 (27.9) |  | 27 (6.9) |  | 114 (28.9) |  | 28 (7.1) |  |
| Other | 1338 (17.2) | 735 (54.9) |  | 398 (29.7) |  | 283 (21.2) |  | 71 (5.3) |  | 299 (22.3) |  | 74 (5.5) |  |
| **Self-perceived health (n=7762)** |  |  |  |  |  |  |  |  |  |  |  |  |  |
| Very good | 2235 (28.8) | 1574 (70.4) | <0.001 | 877 (39.2) | <0.001 | 660 (29.5) | <0.001 | 182 (8.1) | <0.001 | 644 (28.8) | <0.001 | 250 (11.2) | <0.001 |
| Good | 4376 (56.3) | 2572 (58.8) |  | 1254 (28.7) |  | 998 (22.8) |  | 265 (6.1) |  | 1103 (25.2) |  | 362 (8.3) |  |
| Medium | 1078 (13.9) | 482 (44.7) |  | 218 (20.2) |  | 197 (18.3) |  | 41 (3.8) |  | 209 (19.4) |  | 45 (4.2) |  |
| Poor | 62 (0.8) | 23 (37.1) |  | 12 (19.4) |  | 9 (14.5) |  | 0 (0.0) |  | 7 (11.3) |  | 4 (6.5) |  |
| Very poor | 11 (0.1) | 6 (54.5) |  | 5 (45.5) |  | 1 (9.1) |  | 1 (9.1) |  | 1 (9.1) |  | 1 (9.1) |  |
| **Body mass index<sup>d</sup> (n=7654)** |  |  |  |  |  |  |  |  |  |  |  |  |  |
| Under/normal weight (<25kg/m<sup>2</sup>) | 4638 (59.7) | 3033 (65.4) | <0.001 | 1612 (34.8) | <0.001 | 1226 (26.4) | <0.001 | 308 (6.6) | 0.051 | 1308 (28.2) | <0.001 | 438 (9.4) | <0.001 |
| Overweight (25-29.9 kg/m<sup>2</sup>) | 2274 (29.3) | 1254 (55.1) |  | 613 (27.0) |  | 482 (21.2) |  | 149 (6.6) |  | 501 (22.0) |  | 182 (8.0) |  |
| Obesity (≥30 kg/m<sup>2</sup>) | 742 (9.6) | 329 (44.3) |  | 125 (16.8) |  | 136 (18.3) |  | 32 (4.3) |  | 140 (18.9) |  | 38 (5.1) |  |
| **Chronic disease (n=7769)** |  |  |  |  |  |  |  |  |  |  |  |  |  |
| No | 6088 (78.4) | 3849 (63.2) | <0.001 | 2009 (33.0) | <0.001 | 1583 (26.0) | <0.001 | 419 (6.9) | <0.001 | 1619 (26.6) | <0.001 | 542 (8.9) | 0.025 |
| Yes | 1681 (21.6) | 811 (48.2) |  | 359 (21.4) |  | 283 (16.8) |  | 71 (4.2) |  | 345 (20.5) |  | 120 (7.1) |  |
| **Smoking (n=7761)** |  |  |  |  |  |  |  |  |  |  |  |  |  |
| No | 6105 (78.6) | 3785 (62.0) | <0.001 | 1963 (32.2) | <0.001 | 1515 (24.8) | 0.002 | 394 (6.5) | 0.359 | 1600 (26.2) | <0.001 | 543 (8.9) | 0.031 |
| Yes | 1656 (21.3) | 871 (52.6) |  | 403 (24.3) |  | 350 (21.1) |  | 96 (5.8) |  | 362 (21.9) |  | 119 (7.2) |  |
| **Educational level (n=7691)** |  |  |  |  |  |  |  |  |  |  |  |  |  |
| Tertiary | 4003 (51.5) | 2721 (68.0) | <0.001 | 1413 (35.3) | <0.001 | 1093 (27.3) | <0.001 | 335 (8.4) | <0.001 | 1112 (27.8) | <0.001 | 459 (11.5) | <0.001 |
| Upper secondary | 3088 (39.7) | 1661 (53.8) |  | 811 (26.3) |  | 687 (22.2) |  | 142 (4.6) |  | 727 (23.5) |  | 185 (6.0) |  |
| Lower secondary | 600 (7.7) | 229 (38.2) |  | 114 (19.0) |  | 70 (11.7) |  | 11 (1.8) |  | 100 (16.7) |  | 11 (1.8) |  |
| **Household income<sup>e</sup> (n=6927)** |  |  |  |  |  |  |  |  |  |  |  |  |  |
| ≥7000 CHF | 1962 (25.3) | 1367 (69.7) | <0.001 | 738 (37.6) | <0.001 | 542 (27.6) | <0.001 | 173 (8.8) | <0.001 | 531 (27.1) | 0.001 | 266 (13.6) | <0.001 |
| 5000-6999 CHF | 1806 (23.2) | 1157 (64.1) |  | 551 (30.5) |  | 464 (25.7) |  | 126 (7.0) |  | 475 (26.3) |  | 193 (10.7) |  |
| 3000-4999 CHF | 1953 (25.1) | 1065 (54.5) |  | 548 (28.1) |  | 377 (19.3) |  | 93 (4.8) |  | 473 (24.2) |  | 114 (5.8) |  |
| <3000 CHF | 1206 (15.5) | 563 (46.7) |  | 258 (21.4) |  | 238 (19.7) |  | 51 (4.2) |  | 256 (21.2) |  | 38 (3.2) |  |
| **Occupational position (n=5429)** |  |  |  |  |  |  |  |  |  |  |  |  |  |
| Non-manual manager | 1584 (20.4) | 1097 (69.3) | <0.001 | 628 (39.6) | <0.001 | 442 (27.9) | <0.001 | 147 (9.3) | <0.001 | 360 (22.7) | <0.001 | 229 (14.5) | <0.001 |
| Non-manual worker | 2606 (33.5) | 1664 (63.9) |  | 841 (32.3) |  | 671 (25.7) |  | 179 (6.9) |  | 708 (27.2) |  | 220 (8.4) |  |
| Manual self-employed worker | 249 (3.2) | 135 (54.2) |  | 60 (24.1) |  | 43 (17.3) |  | 13 (5.2) |  | 62 (24.9) |  | 14 (5.6) |  |
| Manual worker | 990 (12.7) | 441 (44.5) |  | 260 (26.3) |  | 161 (16.3) |  | 27 (2.7) |  | 155 (15.7) |  | 33 (3.3) |  |

<sup>a</sup> Sport categories: Individual outdoor (running, brisk walking, racing bicycle); Individual with facilities (strength training/weightlifting, swimming, ice-/roller-skating); Racket/martial arts (judo/karate, tennis/badminton, tennis, squash); Group (dance, football, handball, gymnastics); Special (golf, downhill/water skiing, cross-country skiing, diving). Participants can engage in several sports.
<sup>b</sup> Percentages calculated on the total number of participants (7769). Missing categories are not shown, therefore percentages may not add up to 100%. The numbers of observations without missing values are provided in the headline of each variable.
<sup>c</sup> P-values from Chi-Square tests of sport participation adjusted for multiple comparisons with the Bonferroni method
<sup>d</sup> Body mass index categories according to the World Health Organization classification[26]
<sup>e</sup> Gross monthly household income adjusted for the number of people living in the household according to the OECD-modified scale[24]

In line with the descriptive analysis, the RII and SII estimations showed that the higher the socio-economic conditions, the higher the sport participation, whichever the sport category (*P*<0.05; Fig. 1, Supplementary 2-4). For overall sport participation, results of the minimally adjusted model indicated that sport participation was 1.78 times (RII=1.78; 95% CI: 1.64-1.92) and 33 percentage points (SII=0.33; 95% CI: 0.29-0.37), higher among participants with the highest educational level compared to those with the lowest one. These inequalities were lower when adjusted for health-related factors (RII=1.56; 95% CI: 1.45-1.69 and SII=0.26; 95% CI: 0.21-0.30). The same magnitude of results was observed when using household income or occupational position as indicators of socioeconomic circumstances. Relative inequalities were more important for racket/martial arts(RII=3.58; 95% CI: 2.43-5.27) and special sports (RII=4.47; 95% CI: 3.20-6.25) than for group sports (RII=1.76; 95% CI: 1.50-2.06), while the highest absolute inequalities could be observed in the overall sport participation. In analyses stratified by sex, inequalities were similar among men and women, except for group sports where they were higher among women (Supplementary 2-3).

**Fig. 1.**
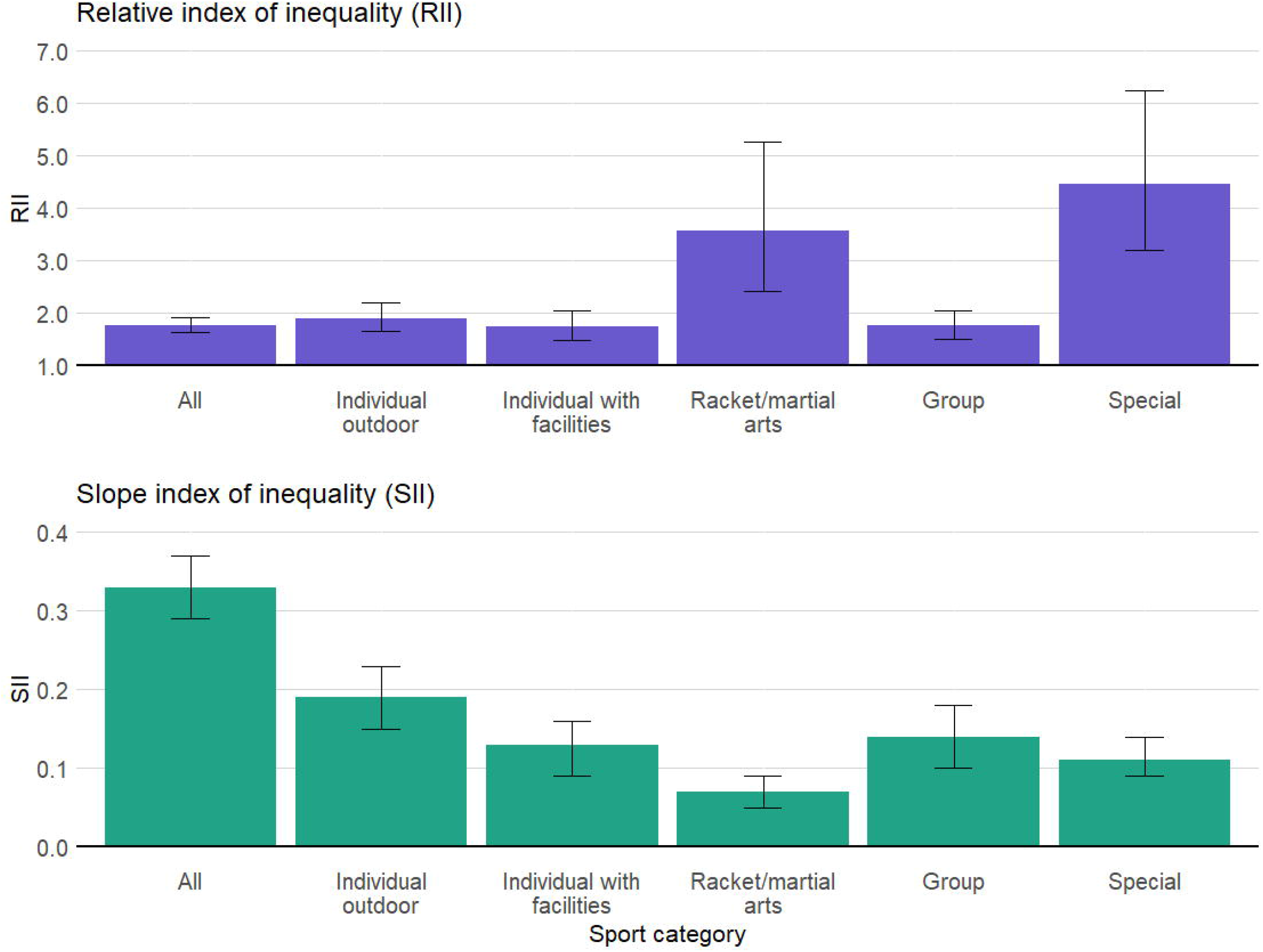
Relative and slope indexes of inequality (RII/SII) and 95% confidence intervals of sport participation among Geneva adults aged 20 to 75 years old, according to the educational level, stratified by sport category. Generalized linear model following a quasi-Poisson distribution for the RII and linear model for the SII. Minimal model, adjusted for age, sex, an interaction between age and sex, and country of birth. Sport categories: Individual outdoor (running, brisk walking, racing bicycle, n=2338); Individual with facilities (strength training/weightlifting, swimming, ice-/roller-skating, n=1866); Racket/martial arts (judo/karate, tennis/badminton, squash, n=488); Group (dance, football, handball, gymnastics, n=1939); Special (golf, downhill/water skiing, cross-country skiing, diving, n=662). N=7689

When stratifying by specific type of activity, for most sports, a higher educational level was associated with higher sport participation. With the minimally adjusted model, sports such as tennis/badminton (RII=4.25; 95% CI: 2.68-6.75) showed high inequalities compared to football (RII=0.68; 95% CI: 0.44-1.07), the only sport where the direction of inequalities seemed reversed, although not significantly (Fig. 2). The same magnitude of results was observed when using the household income and the occupational position as indicators of socioeconomic conditions, apart from substantially higher inequalities in golf participation (Supplementary 5).

**Fig. 2.**
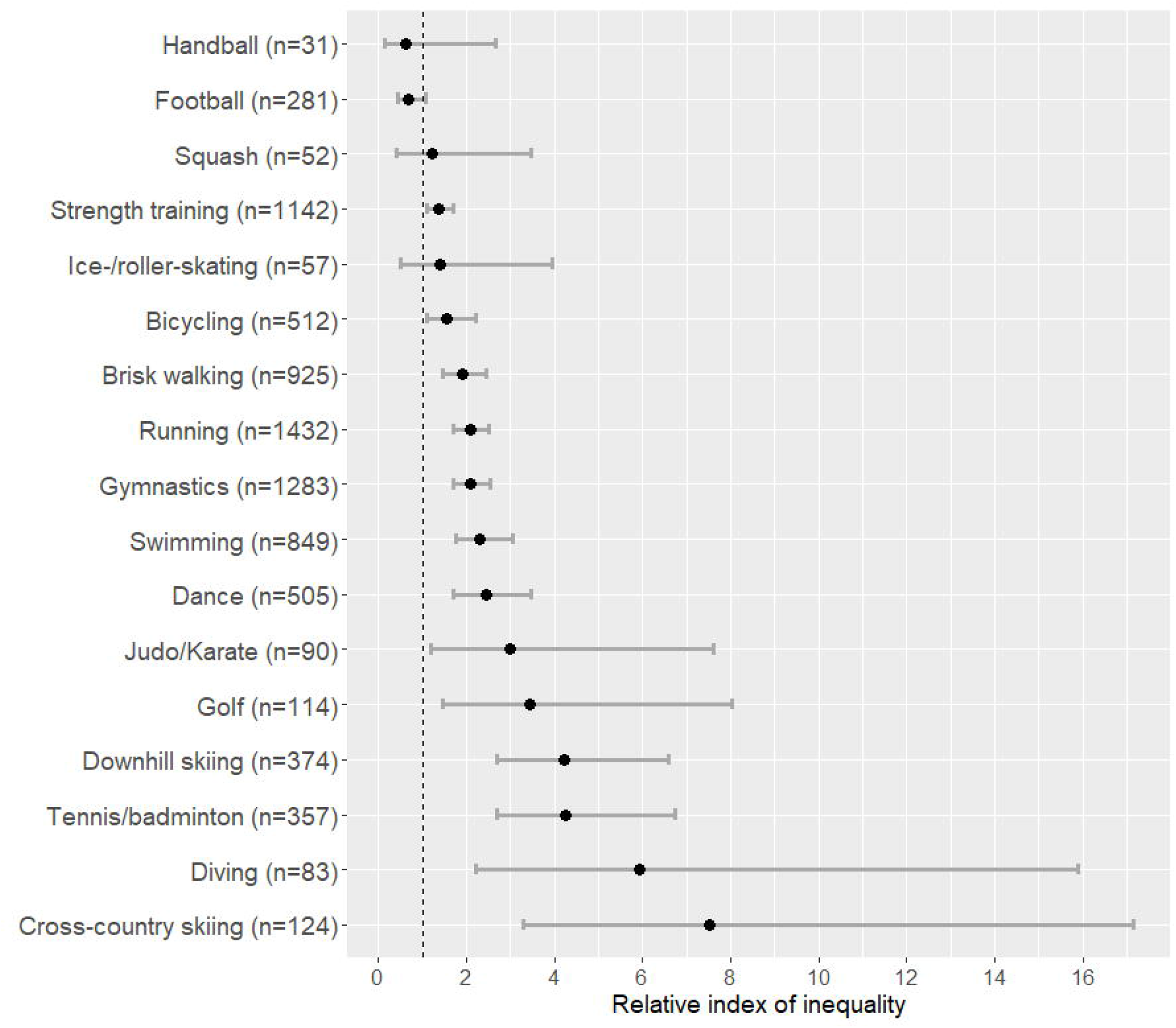
Relative index of inequality (RII) for educational level, stratified by specific sport among Geneva adults aged 20 to 75 years old. Generalized linear model following a quasi-Poisson distribution adjusted for age, sex, an interaction between age and sex, and country of birth. N=7689

Over the 2005-2019 period, for which data was available only for participants aged 35 to 75 years old, the RII and SII showed persistent inequalities. An increasing but non-significant trend could be observed (Fig. 3), for both the yearly change of the RII (exp(β)=1.01; *P=*0.184) and of the SII (β=0.01; *P=*0.099) for education. Absolute income inequalities significantly increased over time (β=0.01; *P=*0.024), especially among women (β=0.02; *P=*0.012; Supplementary 6). Since the comparatively low inequalities observed between 2005 and 2007 (Fig. 3, Supplementary 7) could overly influence these results, a sensitivity analysis was performed by removing these years. When analysing the 2008-2019 period, increasing income inequalities in sport participation were only significant among women (RII: exp(β)=1.03; *P=*0.033 and SII: β=0.02; *P*=0.021; Supplementary 6).

**Fig. 3.**
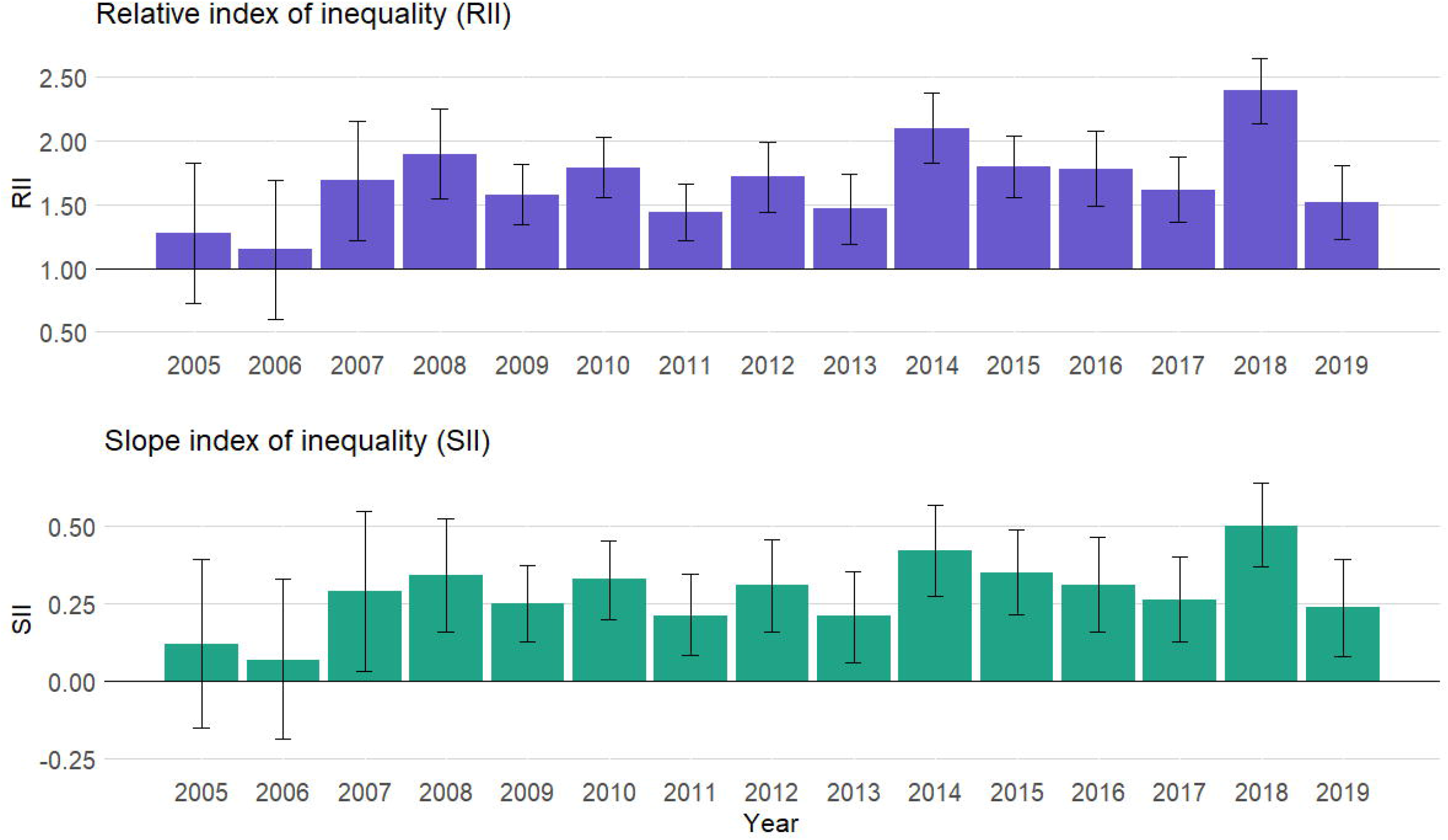
Relative and slope indexes of inequality (RII/SII) and 95% confidence intervals of sport participation according to the educational level, per year, among Geneva adults aged 35 to 75 years old. Generalized linear model following a quasi-Poisson distribution for the RII and linear model for the SII. Minimal model adjusted for sex, age, an interaction between age and sex, and country of birth. N=9711

## 4. DISCUSSION

This study conducted on a randomly selected sample of the Geneva population showed strong socio-economic inequalities in sport participation among adults, with privileged socio-economic conditions being associated with higher sport participation. These inequalities were consistently observed with different socio-economic indicators, as well as after sex and sport stratification. Relative inequalities were higher for racket and special sports than for group sports, while the highest absolute inequalities were observed in the overall sport participation. Income inequalities in sport participation tended to increase over the 2005 to 2019 period.

These results are in line with the existing literature[9–11], and more specifically with other populational studies, that quantified educational inequalities in sporting inactivity in Germany in 2012 (RII=3.4; SII=0.4)[15] and in Sweden in 2014(RII=2.0; SII=0.1) [27]. The high relative inequalities found in the racket, and special sports reflect findings from studies showing that participation in these sports is more common among privileged groups[28,29]. On the other hand, group sport participation seemed to present lower socio-economic inequalities, as previously observed[12,13]. Our results further showed that inequalities in group sport participation were lower among men than women. In fact, sports included in the group sport category (football, handball, gymnastics, and dance) were unequally practised according to gender. Stratifying this category by sex highlighted the higher socio-economic inequalities in gymnastics and dance, which were more practised by women, compared to football and handball, which were popular among men. It shows that analyses by specific sport are relevant to study socio-economic inequalities in sport participation in detail.

Part of the association between socio-economic position and sport participation might be explained by health conditions, as previously observed[30]. Indeed, socio-economic inequalities in sport participation were slightly attenuated when taking health indicators into account. Socio-economic disadvantage is related to poorer health, which may in turn decrease the likelihood of engaging in a sporting activity[30].

Income inequalities in sport participation seemed to increase between 2005 and 2019, especially among women. This is in line with studies finding an increase in socio-economic inequalities in sporting inactivity between 2003 and 2012 in Germany[15] and in health between 1990 and 2010 in Switzerland[31]. Contrary to our results, Galobardes et al.[17] did not observe any trend in socio-economic inequalities in physical inactivity over the 1994-1999 period in Geneva, which suggests that the increasing trend noticed in our study is quite recent. In a context of widening inequalities in income following the 2008 Great Recession[32], it might be that sport activities became relatively less affordable for disadvantaged people. In the wake of the COVID-19 pandemic, it highlights the need for further research to disentangle, and if appropriate to mitigate, the impact of crises on health behaviours.

Because of the positive health impact of leisure-time physical activity, such as sport[2,3,33], our findings are of concern in terms of health inequalities in the population. Our results call for measures encouraging sport participation specifically targeting underprivileged groups, both because they need it the most and because they may not fully benefit from population-level interventions. In Geneva, affordable and quality sport facilities and programs are available to all[34]. Although necessary, such structural measures, especially when relying on wilful behaviours, may not be adapted to limit social inequalities. Indeed, disadvantaged groups may be less aware of the availability of such possibilities, and less likely to make sense of associated recommendations and follow them[35]. Additionally, individuals with lower socioeconomic conditions have been shown to face specific barriers to physical activity such as fatigue, health-related restrictions or financial limitations, their main motivations being enjoyment, health benefits and social interaction or support[36]. To ensure an equitable uptake, targeted interventions should hence aim at reducing these barriers while capitalizing on positive aspects of sport participation. A possibility could be to involve members of underprivileged groups in the delivery of or referral for financially accessible sport sessions[37]. Promoting sporting activities that are already practised in socially disadvantaged communities, while drawing on group interactions and highlighting health benefits could also improve the acceptance of such measures. Finally, it seems important to propose achievable and enjoyable activities that are adapted to the participants needs and abilities beyond their socioeconomic conditions, for instance by also taking their age, gender and health condition into account[36].

This study presents some limitations. Despite the random selection process, individuals with a lower secondary educational level were underrepresented in the sample (7.7% vs 25.4% in the Geneva population)[38]. As health consciousness may be related to a greater interest in epidemiological research, we cannot exclude that underprivileged participants in our study engaged in more sporting activities, than non-participants from similar socio-economic backgrounds. This could lead our results to underestimate socio-economic inequalities in sport participation. On the other hand, social desirability bias related to healthy behaviour was found to be particularly pronounced among privileged individuals[39]. Consequently, they may be more prone than disadvantaged people to over-report sport participation, which in turn could lead to an overestimation of inequalities. The overall prevalence of sport participation may be overestimated, both because of the underrepresentation of disadvantaged individuals who are less likely to engage in a sporting activity, and because of the potential social desirability bias related to the use of self-reported data. Finally, as we assessed sport participation over the previous week, we cannot exclude a seasonal effect in the practice of sporting activities such as skiing or running. However, the effect on our results was thought to be negligible as appointment timing was random over the year.

This study also has major strengths. It relied on a population-based design with a random selection, targeting a large age range. The sizeable sample enabled stratification by sport and sex. Inequalities in sport participation were estimated using three different socio-economic indicators and their evolution over time was measured over 15 years. Finally, we were able to exclude people with reduced mobility to restrict our analyses to individuals able to engage in sporting activities.

## 5. CONCLUSION

This study showed that sport participation is consistently higher among socio-economically advantaged individuals, although the size of inequalities differs according to the type of sport. Group sports showed the lowest relative inequalities, while sizeable inequalities were observed in racket and special sports. These results call for tailored strategies to promote sport participation among socially disadvantaged populations.

## Supporting information

Strobe guidelines

## Data Availability

All data produced in the present study are available upon reasonable request to the authors

## 6. STATEMENTS & DECLARATIONS

### Author Contributions

All authors contributed to the study conception and design. Material preparation and data collection were performed by Giovanni Piumatti. Additional material preparation and analysis were performed by Viviane Richard who also wrote the first draft of the manuscript. All authors critically revised the first draft of the manuscript and approved the final one.

### Ethics approval

The Bus Santé study was approved by the Institute of Ethics Committee of the University of Geneva (ID 10-030R).

### Consent to participate

Written informed consent was obtained from all participants.

## Acknowledgement

We thank the Bus Santé team who collected the data, as well as the participants for their invaluable contribution to the study.

