## Supplementary material for "Socioeconomic inequalities in sport participation: pattern per sport and time trends"

a. Unit of Population Epidemiology, Division of Primary Care Medicine, Geneva University Hospitals, Geneva, Switzerland
b. Fondazione Agnelli, Turin, Italy
c. Department of Health and Community Medicine, Faculty of Medicine, University of Geneva, Geneva, Switzerland
d. Division and Department of Primary Care Medicine, Geneva University Hospitals, Geneva, Switzerland
e. Institute of Global Health, University of Geneva, Geneva, Switzerland.
f. University Center for General Medicine and Public Health, University of Lausanne, Switzerland

CORRESPONDING AUTHOR

Silvia Stringhini

Unité d'épidémiologie populationnelle
Rue Jean-Violette 29
1205 Genève
Switzerland
+41 22 305 58 61

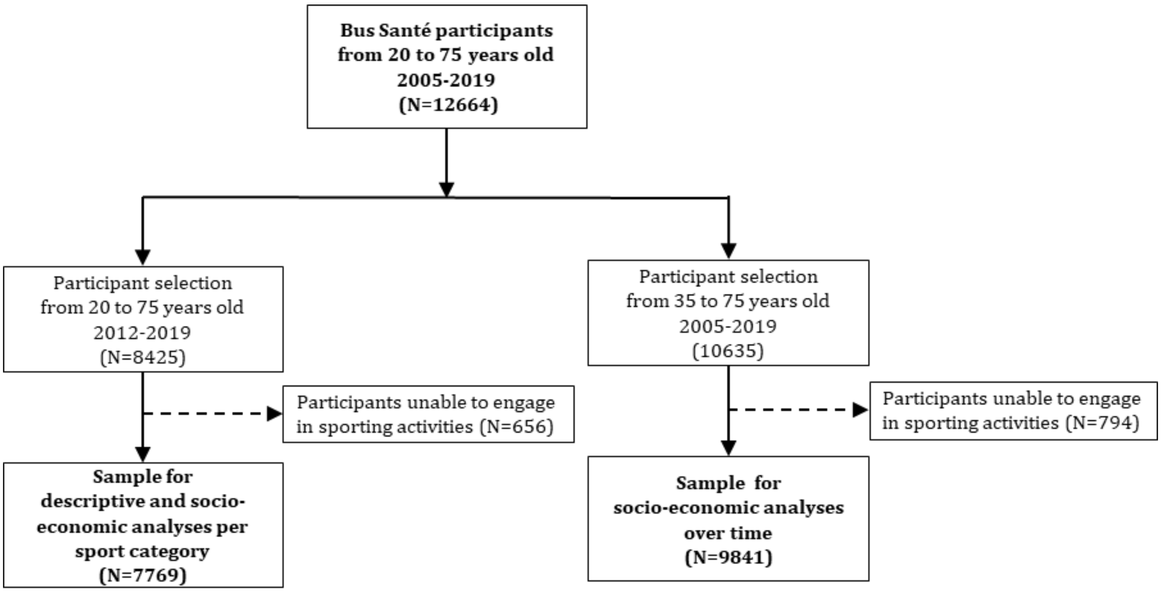


Supplementary 1: Participant's selection process

Supplementary 2: Relative index of inequality (RII) of the educational level, household income and occupational position among adults aged 20 to 75 years old, stratified by sport category and sex

| | All | | | | Men | | | | Women | | | |
|---|---|---|---|---|---|---|---|---|---|---|---|---|
| | Minimally adj.[a] | | Fully adj.[b] | | Minimally adj.[a] | | Fully adj.[b] | | Minimally adj.[a] | | Fully adj.[b] | |
| **Sport category[c]** | **RII** | **CI (95%)** | **RII** | **CI (95%)** | **RII** | **CI (95%)** | **RII** | **CI (95%)** | **RII** | **CI (95%)** | **RII** | **CI (95%)** |
| **All** | | | | | | | | | | | | |
| Educational level | 1.78 | 1.64-1.92 | 1.56 | 1.45-1.69 | 1.77 | 1.59-1.98 | 1.55 | 1.39-1.73 | 1.77 | 1.58-1.98 | 1.56 | 1.39-1.74 |
| Household income | 1.61 | 1.50-1.73 | 1.45 | 1.35-1.55 | 1.65 | 1.49-1.81 | 1.48 | 1.34-1.63 | 1.57 | 1.42-1.74 | 1.40 | 1.27-1.56 |
| Occupational position | 1.60 | 1.47-1.74 | 1.46 | 1.35-1.59 | 1.68 | 1.50-1.88 | 1.54 | 1.38-1.73 | 1.50 | 1.32-1.71 | 1.35 | 1.19-1.54 |
| **Individual outdoor[c]** | | | | | | | | | | | | |
| Educational level | 1.91 | 1.66-2.20 | 1.53 | 1.33-1.77 | 1.95 | 1.62-2.35 | 1.56 | 1.29-1.89 | 1.85 | 1.50-2.29 | 1.48 | 1.19-1.84 |
| Household income | 1.87 | 1.64-2.13 | 1.58 | 1.39-1.80 | 1.89 | 1.60-2.24 | 1.59 | 1.34-1.89 | 1.84 | 1.50-2.25 | 1.53 | 1.25-1.87 |
| Occupational position | 1.77 | 1.52-2.05 | 1.53 | 1.32-1.78 | 2.01 | 1.66-2.44 | 1.75 | 1.44-2.12 | 1.43 | 1.11-1.84 | 1.21 | 0.94-1.57 |
| **Individual with facilities[c]** | | | | | | | | | | | | |
| Educational level | 1.74 | 1.48-2.06 | 1.53 | 1.29-1.82 | 1.88 | 1.50-2.37 | 1.63 | 1.29-2.05 | 1.63 | 1.28-2.06 | 1.46 | 1.14-1.87 |
| Household income | 1.63 | 1.40-1.91 | 1.48 | 1.26-1.73 | 1.55 | 1.26-1.92 | 1.43 | 1.15-1.78 | 1.74 | 1.38-2.19 | 1.56 | 1.23-1.97 |
| Occupational position | 1.90 | 1.58-2.29 | 1.73 | 1.43-2.08 | 1.99 | 1.56-2.54 | 1.83 | 1.43-2.34 | 1.79 | 1.33-2.40 | 1.60 | 1.18-2.17 |
| **Racket/martial arts[c]** | | | | | | | | | | | | |
| Educational level | 3.58 | 2.43-5.27 | 3.09 | 2.09-4.57 | 4.83 | 3.01-7.77 | 4.21 | 2.61-6.78 | 1.83 | 0.94-3.59 | 1.49 | 0.74-2.98 |
| Household income | 2.37 | 1.70-3.31 | 2.08 | 1.48-2.92 | 2.95 | 2.00-4.33 | 2.57 | 1.74-3.81 | 1.29 | 0.66-2.53 | 1.07 | 0.53-2.16 |
| Occupational position | 2.52 | 1.76-3.62 | 2.23 | 1.55-3.22 | 3.17 | 2.11-4.78 | 2.87 | 1.90-4.33 | 1.08 | 0.42-2.75 | 0.78 | 0.29-2.13 |
| **Group[c]** | | | | | | | | | | | | |
| Educational level | 1.76 | 1.50-2.06 | 1.55 | 1.32-1.83 | 1.40 | 1.06-1.83 | 1.24 | 0.94-1.63 | 2.00 | 1.65-2.43 | 1.77 | 1.45-2.16 |
| Household income | 1.38 | 1.19-1.60 | 1.24 | 1.07-1.44 | 1.18 | 0.91-1.52 | 1.04 | 0.80-1.34 | 1.52 | 1.27-1.81 | 1.37 | 1.14-1.64 |
| Occupational position | 1.36 | 1.13-1.64 | 1.26 | 1.04-1.53 | 1.06 | 0.79-1.41 | 0.99 | 0.73-1.33 | 1.66 | 1.31-2.10 | 1.53 | 1.20-1.94 |
| **Special[c]** | | | | | | | | | | | | |
| Educational level | 4.47 | 3.20-6.25 | 3.81 | 2.71-5.38 | 4.78 | 3.07-7.42 | 4.19 | 2.67-6.58 | 4.18 | 2.51-6.97 | 3.41 | 2.01-5.79 |
| Household income | 4.34 | 3.25-5.80 | 3.70 | 2.75-4.98 | 5.23 | 3.58-7.64 | 4.51 | 3.05-6.66 | 3.45 | 2.19-5.42 | 2.94 | 1.85-4.67 |
| Occupational position | 4.06 | 2.88-5.74 | 3.69 | 2.59-5.24 | 4.31 | 2.78-6.70 | 3.99 | 2.55-6.25 | 3.98 | 2.24-7.07 | 3.55 | 1.98-6.38 |

[a] Generalized linear model following a quasi-Poisson distribution, adjusted for age, sex, an interaction between age and sex, and country of birth. [b] Generalized linear model following a quasi-Poisson distribution, adjusted for age, sex, an interaction between age and sex, country of birth, smoking, BMI, self-perceived health and presence of chronic disease. [c] Sport categories: Individual outdoor (running, brisk walking, racing bicycle); Individual with facilities (strength training/weightlifting, swimming, ice-/roller-skating); Racket/martial arts (judo/karate, tennis/badminton, squash); Group (dance, football, handball, gymnastics); Special (golf, downhill/water skiing, cross-country skiing, diving)

Supplementary 3: Slope index of inequality (SII) of the educational level, household income and occupational position among adults aged 20 to 75 years old, stratified by sport category and sex

| | All | | | | Men | | | | Women | | | |
|---|---|---|---|---|---|---|---|---|---|---|---|---|
| | Minimally adj.[a] | | Fully adj.[b] | | Minimally adj.[a] | | Fully adj.[b] | | Minimally adj.[a] | | Fully adj.[b] | |
| **Sport category[c]** | **SII** | **CI (95%)** | **SII** | **CI (95%)** | **SII** | **CI (95%)** | **SII** | **CI (95%)** | **SII** | **CI (95%)** | **SII** | **CI (95%)** |
| **All** | | | | | | | | | | | | |
| Educational level | 0.33 | 0.29-0.37 | 0.26 | 0.21-0.30 | 0.33 | 0.27-0.40 | 0.26 | 0.19-0.32 | 0.33 | 0.26-0.39 | 0.26 | 0.19-0.32 |
| Household income | 0.28 | 0.24-0.32 | 0.22 | 0.18-0.26 | 0.30 | 0.24-0.36 | 0.24 | 0.18-0.30 | 0.27 | 0.21-0.32 | 0.20 | 0.14-0.26 |
| Occupational position | 0.28 | 0.23-0.33 | 0.23 | 0.18-0.28 | 0.32 | 0.25-0.38 | 0.27 | 0.20-0.33 | 0.24 | 0.16-0.31 | 0.18 | 0.10-0.25 |
| **Individual outdoor[c]** | | | | | | | | | | | | |
| Educational level | 0.19 | 0.15-0.23 | 0.13 | 0.08-0.17 | 0.22 | 0.16-0.27 | 0.14 | 0.08-0.20 | 0.16 | 0.11-0.21 | 0.11 | 0.05-0.16 |
| Household income | 0.19 | 0.15-0.23 | 0.14 | 0.10-0.18 | 0.21 | 0.16-0.27 | 0.16 | 0.10-0.22 | 0.16 | 0.11-0.21 | 0.11 | 0.06-0.17 |
| Occupational position | 0.19 | 0.14-0.23 | 0.14 | 0.09-0.19 | 0.25 | 0.18-0.32 | 0.20 | 0.13-0.27 | 0.10 | 0.03-0.17 | 0.06 | -0.02-0.13 |
| **Individual with facilities[c]** | | | | | | | | | | | | |
| Educational level | 0.13 | 0.09-0.16 | 0.10 | 0.06-0.13 | 0.15 | 0.09-0.20 | 0.11 | 0.06-0.17 | 0.11 | 0.05-0.16 | 0.08 | 0.03-0.14 |
| Household income | 0.11 | 0.08-0.15 | 0.09 | 0.05-0.13 | 0.10 | 0.05-0.16 | 0.09 | 0.03-0.14 | 0.12 | 0.07-0.17 | 0.10 | 0.05-0.15 |
| Occupational position | 0.15 | 0.10-0.19 | 0.12 | 0.08-0.17 | 0.17 | 0.11-0.23 | 0.14 | 0.08-0.20 | 0.12 | 0.06-0.19 | 0.10 | 0.03-0.16 |
| **Racket/martial arts[c]** | | | | | | | | | | | | |
| Educational level | 0.07 | 0.05-0.09 | 0.06 | 0.04-0.09 | 0.13 | 0.09-0.16 | 0.12 | 0.08-0.15 | 0.02 | 0.00-0.04 | 0.01 | -0.01-0.04 |
| Household income | 0.05 | 0.03-0.07 | 0.05 | 0.02-0.07 | 0.10 | 0.06-0.13 | 0.09 | 0.05-0.12 | 0.01 | -0.01-0.03 | 0.00 | -0.02-0.03 |
| Occupational position | 0.06 | 0.04-0.09 | 0.05 | 0.03-0.08 | 0.11 | 0.07-0.15 | 0.10 | 0.06-0.14 | 0.00 | -0.03-0.03 | -0.01 | -0.04-0.02 |
| **Group[c]** | | | | | | | | | | | | |
| Educational level | 0.14 | 0.10-0.18 | 0.11 | 0.07-0.15 | 0.06 | 0.01-0.11 | 0.04 | -0.01-0.09 | 0.21 | 0.15-0.26 | 0.17 | 0.11-0.23 |
| Household income | 0.08 | 0.04-0.12 | 0.05 | 0.02-0.09 | 0.03 | -0.02-0.08 | 0.01 | -0.04-0.06 | 0.13 | 0.07-0.19 | 0.10 | 0.04-0.16 |
| Occupational position | 0.07 | 0.03-0.11 | 0.05 | 0.01-0.10 | 0.01 | -0.04-0.06 | 0.00 | -0.06-0.05 | 0.14 | 0.08-0.21 | 0.12 | 0.05-0.19 |
| **Special[c]** | | | | | | | | | | | | |
| Educational level | 0.11 | 0.09-0.14 | 0.10 | 0.08-0.12 | 0.14 | 0.10-0.18 | 0.13 | 0.09-0.17 | 0.09 | 0.06-0.11 | 0.07 | 0.04-0.10 |
| Household income | 0.12 | 0.10-0.15 | 0.11 | 0.09-0.13 | 0.16 | 0.13-0.20 | 0.15 | 0.11-0.19 | 0.08 | 0.05-0.11 | 0.07 | 0.04-0.10 |
| Occupational position | 0.12 | 0.09-0.15 | 0.11 | 0.08-0.14 | 0.15 | 0.11-0.20 | 0.14 | 0.10-0.19 | 0.08 | 0.05-0.12 | 0.08 | 0.04-0.12 |

[a] Linear model adjusted for age, sex, an interaction between age and sex, and country of birth. [b] Linear model adjusted for age, sex, an interaction between age and sex, country of birth, smoking, BMI, self-perceived health and presence of chronic disease. [c] Sport categories: Individual outdoor (running, brisk walking, racing bicycle); Individual with facilities (strength training/weightlifting, swimming, ice-/roller-skating); Racket/martial arts (judo/karate, tennis/badminton, squash); Group (dance, football, handball, gymnastics); Special (golf, downhill/water skiing, cross-country skiing, diving)

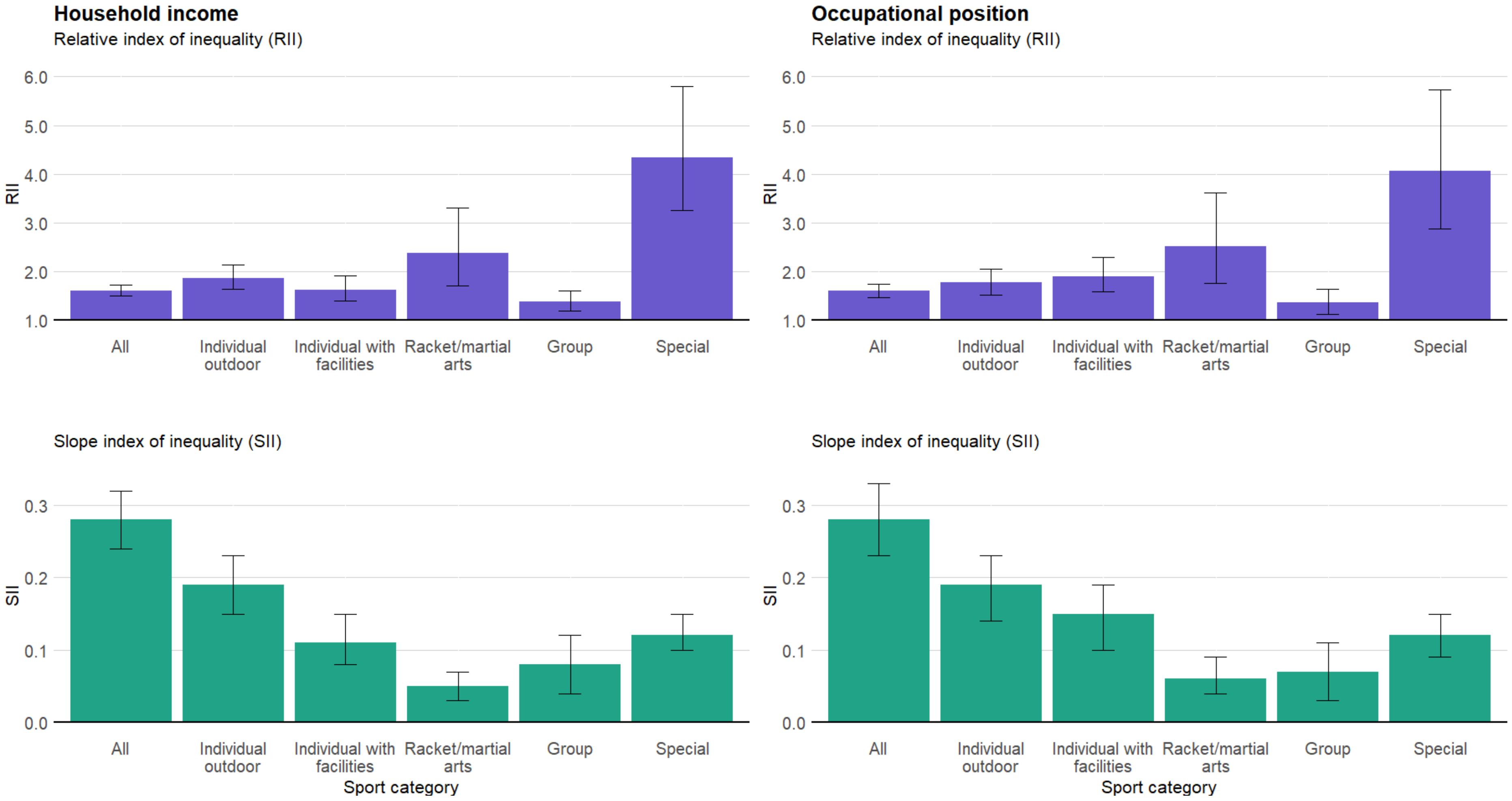


Supplementary 4: Relative and slope indexes of inequality (RII/SII) and 95% confidence intervals of sport participation among Geneva adults aged 20 to 75 years old stratified by sport category. Generalized linear model following a quasi-Poisson distribution for the RII and linear model for the SII. Minimal model, adjusted for age, sex, an interaction between age and sex, and country of birth. Sport categories: Individual outdoor (running, brisk walking, racing bicycle); Individual with facilities (strength training/weightlifting, swimming, ice-/roller-skating); Racket/martial arts (judo/karate, tennis/badminton, squash); Group (dance, football, handball, gymnastics); Special (golf, downhill/water skiing, cross-country skiing, diving). N=6925 for household income and n=5428 for occupational position.

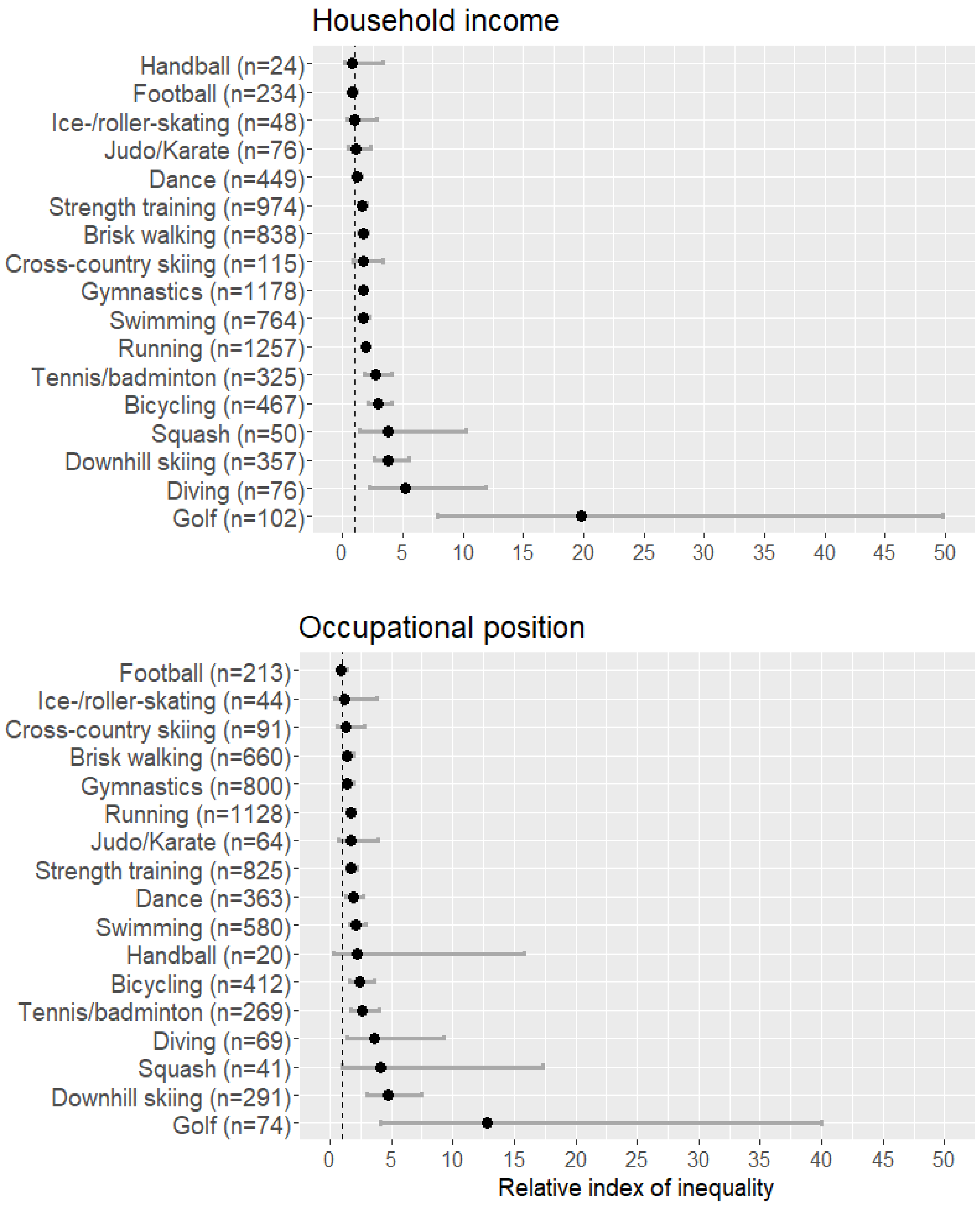


Supplementary 5: Relative index of inequality (RII), stratified by specific sport among Geneva adults aged 20 to 75 years old. Generalized linear model following a quasi-Poisson distribution adjusted for age, sex, an interaction between age and sex, and country of birth. N=6925 for household income and n=5428 for occupational position.

Supplementary 6: Yearly change of the relative and slope index of inequality (RII/SII) of sport participation from 2005 to 2019 among adults aged 35 to 75 years old, and sensitivity analysis from 2008 to 2019

| | **All** | | | | **Men** | | | | **Women** | | | |
|---|---|---|---|---|---|---|---|---|---|---|---|---|
| | **Minimally adj.[c]** | | **Fully adj.[d]** | | **Minimally adj.[c]** | | **Fully adj.[d]** | | **Minimally adj.[c]** | | **Fully adj.[d]** | |
| | **β** | **p-value** | **β** | **p-value** | **β** | **p-value** | **β** | **p-value** | **β** | **p-value** | **β** | **p-value** |
| **2005 - 2019** | | | | | | | | | | | | |
| **Relative index of inequality [a]** | | | | | | | | | | | | |
| Educational level | 1.01 | 0.184 | 1.02 | 0.119 | 1.02 | 0.180 | 1.02 | 0.348 | 1.01 | 0.421 | 1.01 | 0.184 |
| Household income | 1.02 | 0.064 | 1.01 | 0.281 | 1.01 | 0.618 | 0.99 | 0.444 | 1.03 | 0.028 | 1.02 | 0.064 |
| Occupational position | 1.01 | 0.412 | 1.00 | 0.794 | 1.01 | 0.454 | 1.00 | 0.913 | 1.01 | 0.665 | 1.01 | 0.412 |
| **Slope index of inequality [b]** | | | | | | | | | | | | |
| Educational level | 0.01 | 0.099 | 0.01 | 0.067 | 0.01 | 0.169 | 0.01 | 0.242 | 0.01 | 0.317 | 0.01 | 0.099 |
| Household income | 0.01 | 0.024 | 0.01 | 0.148 | 0.01 | 0.405 | 0.00 | 0.684 | 0.02 | 0.012 | 0.01 | 0.024 |
| Occupational position | 0.01 | 0.248 | 0.00 | 0.587 | 0.01 | 0.309 | 0.00 | 0.849 | 0.01 | 0.589 | 0.01 | 0.248 |
| **2008-2019** | | | | | | | | | | | | |
| **Relative index of inequality [a]** | | | | | | | | | | | | |
| Educational level | 1.01 | 0.404 | 1.01 | 0.409 | 1.01 | 0.632 | 1.00 | 0.847 | 1.01 | 0.380 | 1.02 | 0.281 |
| Household income | 1.01 | 0.167 | 1.01 | 0.442 | 1.00 | 0.999 | 0.98 | 0.394 | 1.03 | 0.033 | 1.03 | 0.045 |
| Occupational position | 1.01 | 0.336 | 1.00 | 0.748 | 1.01 | 0.585 | 1.00 | 0.911 | 1.02 | 0.379 | 1.00 | 0.829 |
| **Slope index of inequality [b]** | | | | | | | | | | | | |
| Educational level | 0.01 | 0.330 | 0.01 | 0.317 | 0.00 | 0.681 | 0.00 | 0.777 | 0.01 | 0.308 | 0.01 | 0.254 |
| Household income | 0.01 | 0.097 | 0.01 | 0.283 | 0.00 | 0.802 | -0.01 | 0.573 | 0.02 | 0.021 | 0.02 | 0.032 |
| Occupational position | 0.01 | 0.244 | 0.00 | 0.587 | 0.01 | 0.460 | 0.00 | 0.732 | 0.01 | 0.332 | 0.00 | 0.801 |

β : yearly change of the RII/SII (exponential of the coefficient of the socioeconomic indicator x year term for the RII; coefficient of the socioeconomic indicator x year term for the SII)

[a] Generalized linear model following a quasi-Poisson distribution; [b] Linear model; [c] Adjusted for socioeconomic indicator, age, sex, country of birth, year, and interactions: age x sex, socioeconomic indicator x year, age x year, sex x year, country of birth x year; [d] Adjusted for socioeconomic indicator, age, sex, country of birth, smoking, BMI, self-perceived health, presence of chronic disease and interactions: year, age x sex, socioeconomic indicator x year, age x year, sex x year, country of birth x year, smoking x year, BMI x year, self-perceived health x year, presence of chronic disease x year.

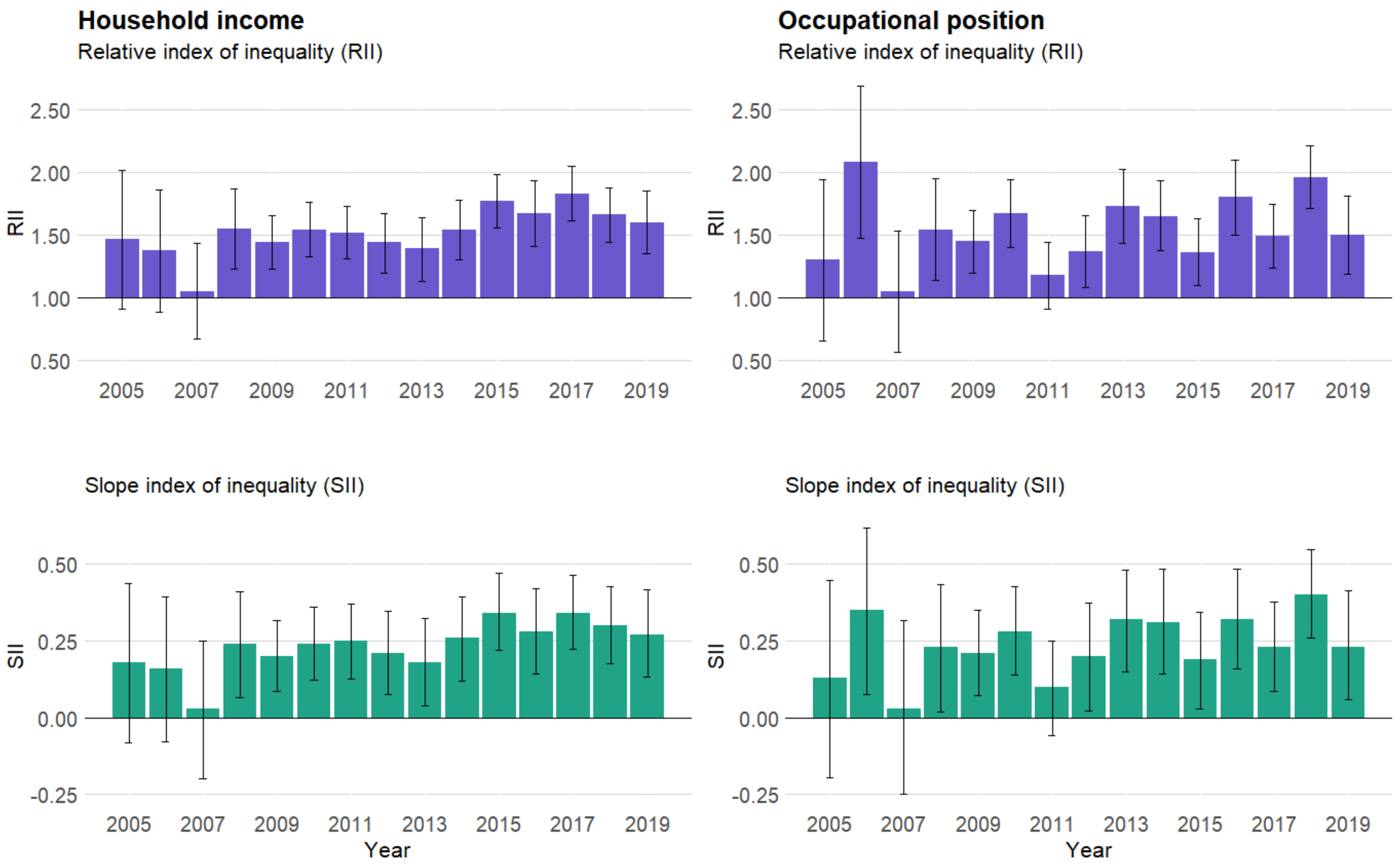


Supplementary 7: Relative and slope indexes of inequality (RII/SII) and 95% confidence intervals of sport participation, per year, among Geneva adults aged 35 to 75 years old. Generalized linear model following a quasi-Poisson distribution for the RII and linear model for the SII. Minimal model adjusted for sex, age, an interaction between age and sex, and country of birth. N=9251 for household income and n=6967 for occupational position.
